# A rights-based intervention integrating social work and ophthalmic care for people experiencing or at risk of homelessness

**DOI:** 10.64898/2026.04.22.26351525

**Authors:** Abolfazl Hassani, Kate Pecar, Mary Soliman, Phil Bunyon, Chris Ellinger, Grace Tulysewskid, Jo Croft, Cesar Carillo, Gemma Wewegam, Sharon du Plessis-Schneider, Jose J. Estevez

## Abstract

**Background:** Individuals experiencing or at risk of homelessness face substantial barriers to preventive eye care that are poorly addressed by standard service models. Interdisciplinary optometry–social work collaboration offers a rights-based approach to improving engagement and continuity of care.

**Methods:** A convergent mixed-methods study was conducted between February and August 2024 at a multidisciplinary community centre. Clients experiencing or at risk of homelessness received integrated optometry and social work assessment and were prioritised as high, medium, or low based on combined clinical and social risk. Social work follow-up was guided by the Triple Mandate and W-Questions framework. Quantitative data were summarised using mean (SD), median [IQR], or n (%). Qualitative case notes were analysed using content analysis with inductive coding and secondary review for consistency.

**Results:** A total of 165 clients had priority categories coded (high: 68; medium: 47; low: 154). Demographic data were available for 132 clients (60% male; mean age 49.5 years [SD 16]); 27% had not completed high school, 89% reported weekly income below AUD 1000, and 28% had vision impairment. Two hundred forty-five case-note entries were consolidated into 146 unique records. SMS (46%) and phone calls (38%) were the most documented contact methods, although only 21% of calls were answered; missed calls (13%) and disconnected numbers (7%) were common. Multi-modal contact was more frequently documented for higher-priority clients. Appointment assistance was the most recorded facilitator (71%), while rights-based supports, including interpreter and transport assistance, were infrequently documented (≤5%). Qualitative analysis identified unstable communication, reliance on informal supports, and service fragmentation as key influences on recall outcomes.

**Conclusion:** This study supports an interdisciplinary, rights-based optometry–social work model to address barriers to preventive eye care among people experiencing or at risk of homelessness. Embedding structured handovers and tiered recall processes within community-based services may strengthen continuity and accountability for high-priority clients. Future implementation should evaluate outcomes related to equity of reach, service integration, and sustained engagement in care.

## Introduction

People experiencing or at risk of homelessness who have poor vision face higher risks of injury and reduced capacity to engage with services, education, and employment, which in turn undermines housing stability and reinforces cycles of disadvantage.^1^ Globally, a review of 23 studies reported that one in four individuals experiencing homelessness had functional visual impairment as a consequence of higher rates of uncorrected refractive error, cataract, glaucoma, and diabetic retinopathy compared to housed individuals of similar age.^2^ Although equitable healthcare is a universally recognised right, access to preventive eye care remains limited for many experiencing or at risk of homelessness, partly due to structural barriers that the eye care discipline cannot overcome without collaboration from broader healthy and social support systems.^3^ Structural factors such as transportation difficulties, inflexible scheduling, stigma, and fragmented care systems hinder timely and effective service delivery.^4–6^ Service mapping studies on outreach models such as community-based screenings have identified persistent gaps in coordinated care pathways and highlighted the importance of non-clinical supports, including care navigation, culturally appropriate engagement strategies, and integration with other health and social services to sustain participation in eye health programs.^7^

Outreach care models are typically siloed and struggle to address the intersection of eye healthcare needs and broader social determinants, particularly for individuals experiencing or at risk of homelessness.^8^ A more potentially effective solution lies in interdisciplinary collaboration, where eye care clinicians (optometry or ophthalmology), social work and other disciplines collaborate to deliver coordinated and holistic services. Interdisciplinary collaboration integrates different disciplinary knowledge bases to address complex health and social issues.^9^ This approach also provides opportunities for interprofessional education, equipping students across disciplines with the collaborative practice skills needed to provide coordinated, patient-centred care.^10^ Evidence demonstrates that such integrated care models may improve service access, clinical outcomes, and patient experience among priority populations.^11,12^

The current study aims to explore an interdisciplinary collaboration approach that integrates optometry and social work services within a rights-based framework, asserting that access to preventive eye care is a fundamental human right that requires both clinical expertise and the removal of systemic barriers.^13^ This assertion finds theoretical grounding in Article 12 of the International Covenant on Economic, Social and Cultural Rights, which recognises “*the right of everyone to the enjoyment of the highest attainable standard of physical and mental health*”.^14^ Social work within a rights-based framework refers to practice guided by human dignity and social justice, which recognises human rights as the foundation for addressing social problems and requires coordinated action across individual and organisational levels (within service systems such as the community centre), and ultimately across system-level factors such as funding arrangements, policies, and regulatory frameworks.^15–17^ Drawing on the Triple Mandate and the W-Questions frameworks for structured intervention planning, social work conceptualises accountability to clients, institutions, and professional ethics while using the W-Questions to structure assessment and follow-up. The study outlines the approach and identifies key themes within an integrated social work–optometry model to enhance access, utilisation, and outcomes.^18,19^ Evaluating the implementation of this collaborative model addresses a gap in the literature on integrated service delivery for populations experiencing systemic inequities. Additionally, it contributes evidence supporting rights-based healthcare approaches.

## Methods

### Study design

A convergent mixed-methods approach was used to evaluate an interdisciplinary collaborative eye care and social work service for individuals experiencing or at risk of homelessness. Quantitative and qualitative data collected over seven months of the optometry–social work model were analysed concurrently, and regular team meetings were held to discuss qualitative findings on client barriers and engagement, which informed small operational changes to the clinic process and referral pathways. All consecutive clients attending the on-site eye clinic during the study period were eligible. We excluded duplicate encounters for the same client within the analysis window, retaining only the first complete assessment. For descriptive items (e.g., education, income, acuity), we used available-case denominators, which are reported alongside results. **Figure 1** provides an overview of client inclusion and data flow across the quantitative and qualitative components of the study.

**Figure 1:**
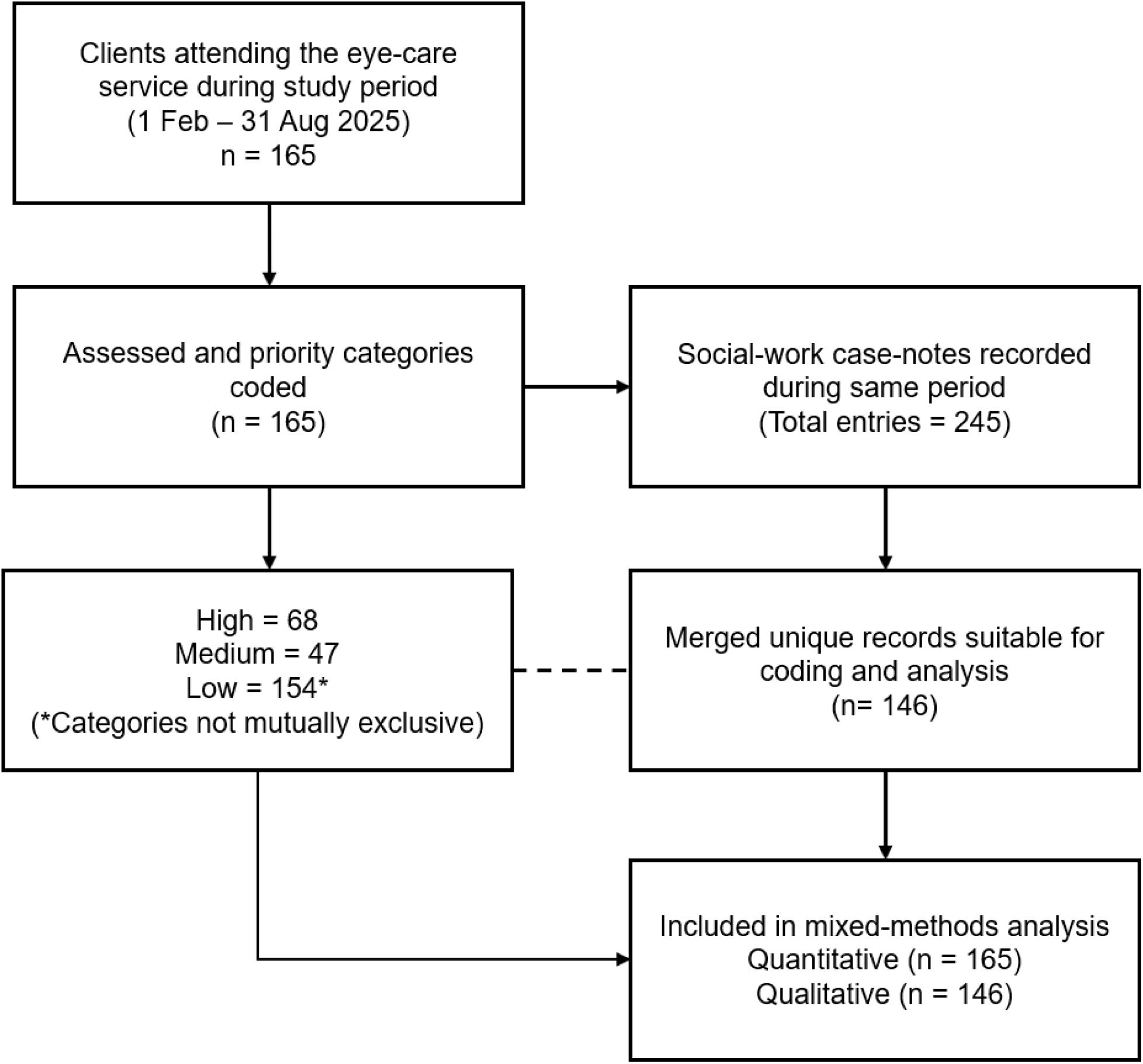
Flowchart showing clients and records included in the quantitative (clinical) and qualitativ (social work) components of the study. The dashed line indicates integration of datasets for mixed-methods analysis. *Categories not mutually exclusive; n = number of clients or records*.

### Study setting

The setting was a community centre in a major Australian city offering a range of co-located multidisciplinary services for individuals experiencing or at risk of homelessness. Services include primary healthcare, mental health support, legal advice, income support and disability advocacy, and health clinics in eye care, dentistry, and physiotherapy. The centre also provides food and transport vouchers, free meals, and weather-contingent emergency shelter. A dedicated program supports social participation and helps individuals access long-term housing and healthcare.

### Structure of the intervention

The intervention was developed via the W-Questions framework to address barriers to care.^19^ The integrated model was co-designed by the community centre, university-based optometry and social work programs, hospital-based ophthalmology services, and partner organisations. Final-year optometry students from Flinders University conducted comprehensive eye examinations under the supervision of an Australian-registered optometrist. These included LogMAR visual acuity assessment, retinoscopy, subjective refraction, intraocular pressure measurement, slit-lamp evaluation of anterior and posterior segments, and fundus photography. Ancillary investigations, such as optical coherence tomography and visual field testing, were conducted when clinically indicated. Prescription spectacles were dispensed at no cost through industry partners’ support. Individuals requiring tertiary-level care were referred to the local ophthalmology department at a public hospital that was a project partner. Post-discharge follow-up appointments were primarily arranged by the social work team, in coordination with the supervising optometrist. Presenting binocular acuity in the better eye worse than 0.3 logMAR defined the presence of vision impairment.

Social work students, under the supervision of a qualified social worker, assessed social and emotional well-being using discipline-specific approaches informed by structured problem and resource analysis tool, the W-Questions framework.^15^ Assessments followed the W-Questions domains (who, what, why, how, with whom, which structural or systemic factors), identifying who was affected and what barriers limited engagement across housing, income, identification, language, transport, employment, literacy, and social connections. Informed by the Triple Mandate, which requires accountability to clients, the community centre, and the profession’s ethical-scientific standards, students coordinated supports, arranged follow-ups, assisted with identification, and liaised with services to strengthen care continuity.^18^ Where necessary, both social worker and students accompanied clients to external appointments and liaised with hospital departments.

### Clients and priority assessment

Integrated case conferencing and a shared client record enabled real-time interdisciplinary communication between the optometry and social work teams. At the conclusion of each eye examination, the optometrist assigned a single priority category (high, medium, or low) to each client. Priority was assigned at the individual client level, based on the clinician’s whole-of-person assessment available demographic, clinical, and social information at the time of assessment. Key clinical considerations included visual impairment, age (i.e., over 50 years old or minors), a documented chronic systemic disease with known ocular complications (i.e., diabetes), and clinical indicators of elevated risk for ocular morbidity identified during examination. Relevant social factors, such as duration of homelessness or housing insecurity and the availability of existing support systems, were also considered where known. The assigned priority level was used to support care coordination and case management discussions with the social work team, assisting in the timely identification of clients requiring more urgent follow-up or support. Priority assignment functioned as a service-level coordination tool rather than a validated outcome measure and was analysed descriptively to characterise patterns of urgency and follow-up.

### Data analysis

Qualitative data were summarised using qualitative content analysis.^20^ Categories were deductively defined by the researchers based on established procedures for case note documentation, including the reason for contact, the method of communication, and the content of interactions, and codes were developed inductively, following a data-driven approach, to identify and capture patterns within the data. One researcher (KP) coded all records; a second researcher (JE) reviewed a sample for concordance; disagreements were resolved by discussion. Continuous variables were summarised as mean (SD) or median (IQR) (as appropriate) and categorical variables as n (%). Records with unclear/insufficient detail for a category were coded “unclear/unspecified” and excluded from category percentages but retained in overall counts. Data were de-identified before analysis and stored on secure, access-controlled servers. Quantitative data were analysed in STATA v.18 (StataCorp, Texas).

### Ethics

Ethical approval for this study was granted by the Central Adelaide Local Health Network Human Research Ethics Committee (HREC#17661). Additional letters of support were provided by the major public hospital’s ophthalmology department, the community centre, and the university. All participants provided written informed consent before study procedures. The study was conducted in accordance with the National Statement on Ethical Conduct in Human Research and the principles of the Declaration of Helsinki.

## Results

Between 1 February and 31 August 2024, a total of 165 clients attended the eye-care service during the study period. Clinical and service-related categories for each client were coded into priority levels to describe the distribution of urgency across presenting categories. Of the 132 clients who reported demographic information, 60% (n=79/132) identified as male and 40% (n=53/132) self-identified as Caucasian. The mean age was 49.5 years (SD 16). The average duration of homelessness or housing instability was 28.1 months (SD 34). Among the 132 clients with available education and income data, 27% (36/132) had not completed high school and 89% (117/132) reported a weekly income below $1,000 AUD. Vision impairment was present in 28% of clients with available data (n=37/132). Available-case denominators are reported for variables with missing data.

Overall, 32 priority assignments were classified as high, 30 as medium, and 103 as low. Table 1 summarises priority assignment across clinical categories. Of 245 total case notes, 146 unique merged social work records were suitable for coding and analysis, reflecting consolidation of multiple entries per client (Table 2). The median number of entries per client was 2 (range 1–11). Short message service (SMS) and phone were the most frequent contact methods (46% and 38% of coded entries, respectively). Across outgoing phone calls, 21% were answered, 13% had no answer, 7% reached disconnected services, and 42% had unclear or unspecified outcomes. Most calls were directed to the client (84%), with smaller proportions to social workers (8%) and external providers (6%). The most common topics were booking an appointment (43%), reminders for eye care (24–26%), and post-visit feedback (4%). Documented facilitators of engagement included appointment assistance (71%), interpretation or translation support (5%), and involvement of a support person (5%). Challenges included appointments not booked at the point of care (82% of challenge entries) and delays in spectacle delivery (18%).

**Table 1.**
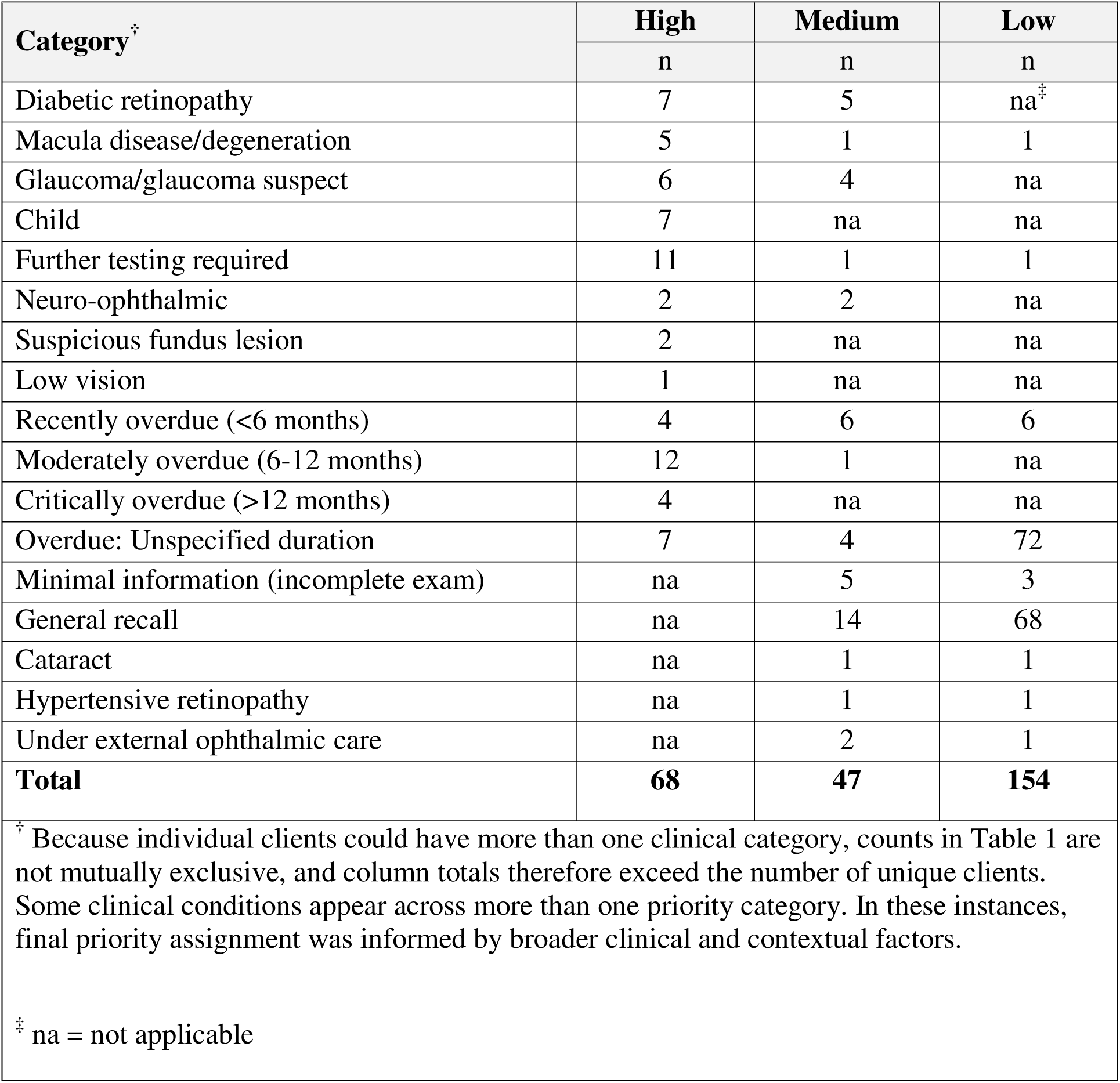
Priority assignment (n=165)

**Table 2.**
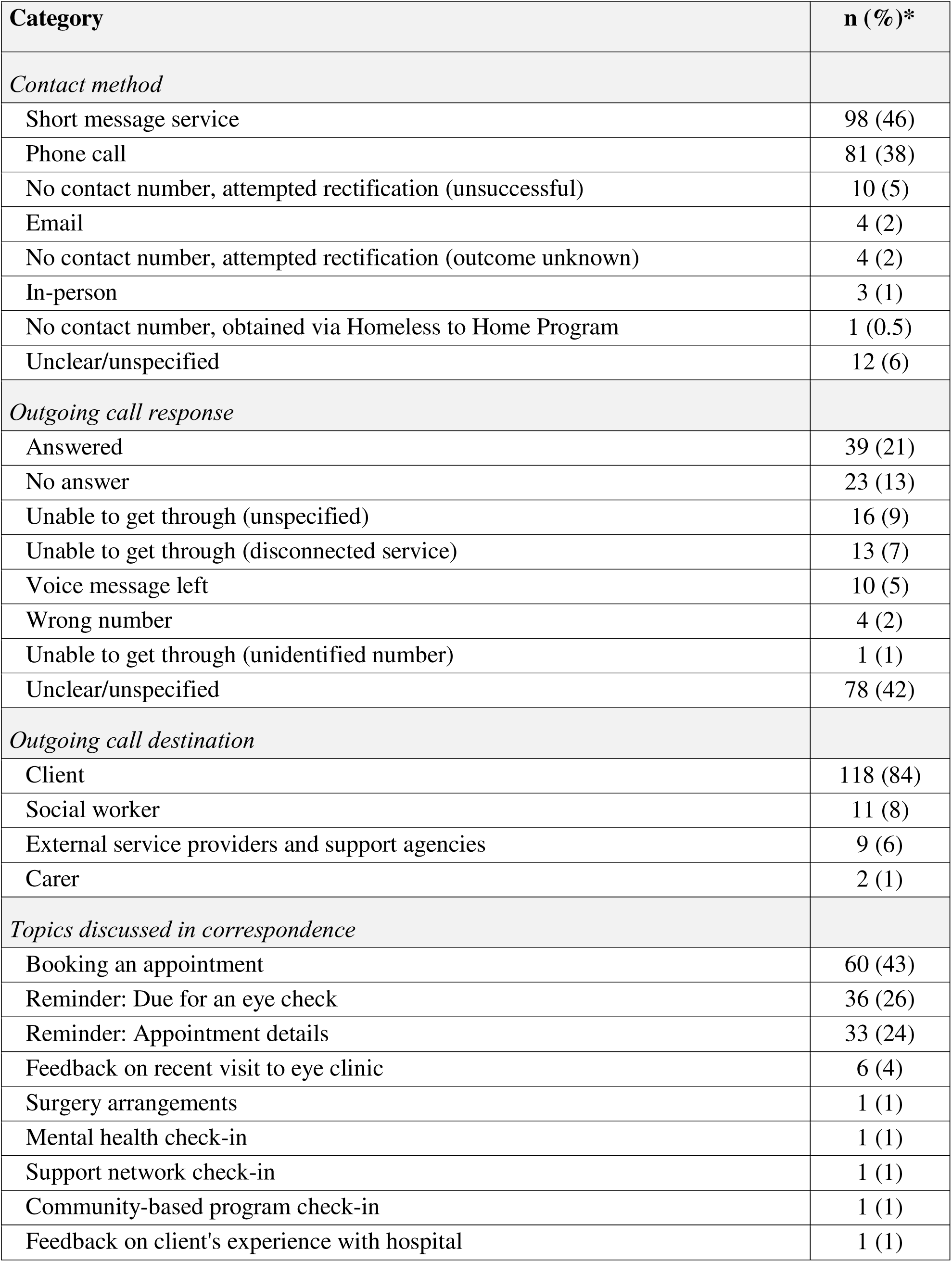
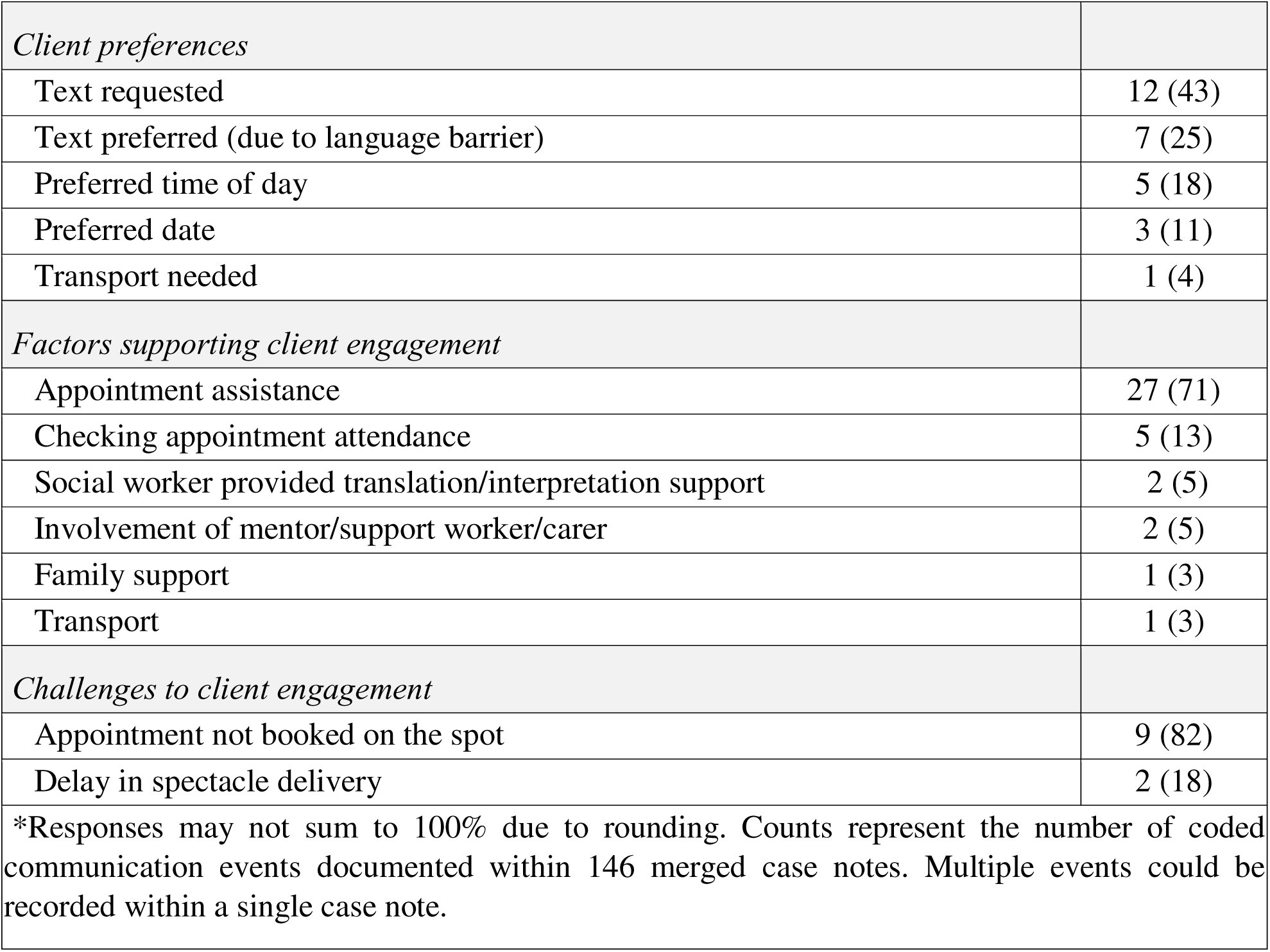
Client communication, engagement efforts and key themes in social worker case notes.

## Discussion

This study evaluated an interdisciplinary, rights-based eye care model integrating optometry and social work to address barriers to preventive eye care among individuals experiencing or at risk of homelessness. Guided by the Triple Mandate and W-Questions frameworks, the intervention sought to uphold the ethical right to equitable care, align with organisational service delivery objectives, and promote client wellbeing through structured priority grading, follow-up, and support.^15,18^ By combining clinical and social perspectives, the model demonstrated how coordinated care can reduce inequities that neither discipline can address in isolation.

A total of 165 clients were prioritised, and 146 social case notes were analysed to identify service delivery patterns and recall outcomes. The main contact methods were SMS (46%) and phone calls (38%), yet only 21% of calls were answered, 13% went unanswered, 7% reached disconnected numbers, and 42% lacked a recorded outcome. These findings highlight persistent structural barriers despite coordinated efforts, reflecting well-documented challenges of fragmented recall systems, unstable communication channels, and limited resourcing in services supporting people experiencing homelessness.^5^ Documented facilitators such as appointment assistance, transport, and interpreter support were offset by recurring barriers including unbooked follow-ups and inconsistent documentation. Outcomes of SMS recalls were rarely reported, and reliance on informal interpreting or absent handovers exposed system-level gaps across client, organisational, and professional mandates. Interpreted through the Triple Mandate and W-Questions frameworks,^15,18^ these communication failures indicate misalignments between clients’ right to accessible care (“who is being missed?”), organisational accountability for recall systems (“what processes are failing?”), and the professional obligation to ensure timely follow-up (“why does recall matter?”). Embedding these frameworks into both service design and evaluation can strengthen accountability and responsiveness for clients at the margins of care systems.

The structured priority system linked clinical urgency with social context by assigning high, medium, or low priority levels to clinical and service categories. SMS reminders were commonly used for lower-priority clients, while higher priorities received phone or coordinated follow-up. This approach was consistent with the Royal Australian College of General Practitioners’ Standards, which recommend multiple and escalating contact methods for patients at higher risk.^21^ The model also distinguished between reminders, which encourage attendance, and recalls, which arise from clinical findings and create a duty of follow-up. Linking recall procedures to social context provided a consistent and accountable structure for community and allied health services, addressing known gaps in recall systems for priority populations.

Evidence from primary care shows that SMS reminders can be cost-effective and improve attendance compared with no contact, while telephone calls are generally more effective for high-risk groups with repeated non-attendance.^22–29^ In this study, both methods were applied according to client priority, yet overall contact success remained limited: only 20% of calls were answered, 13% had no response, 7% reached disconnected numbers, and 42% were undocumented. Incomplete recording of call outcomes highlights the need for standardised categories such as “delivered,” “answered,” “no answer,” “disconnected,” or “voicemail” to support accurate monitoring of recall performance. Some clients preferred SMS for language reasons, whereas others could not be reached because of unstable phone access or disconnected services. Limited phone stability, data restrictions, and low digital literacy are well-recognised contributors to digital exclusion and impede continuity of care among hard-to-reach groups.^30^ A systematic review found that SMS reminder systems often fail when clients experience inconsistent device access, limited data, or low digital literacy,30 and a pilot study of veterans experiencing homelessness reported frequent service interruptions that disrupted continuity of care.^31^ These findings indicate that even structured recall systems require adaptable, multi-modal strategies that reflect clients’ communication resources and life circumstances. In this model, SMS was mainly used for lower-priority clients, combined SMS and phone follow-up for medium-priority clients and coordinated social-work or in-person contact for high-priority clients.

Language and cultural access emerged as important structural considerations. Interpreter use was rare (5%), and informal translation was often provided by bilingual staff or support persons, which may have compromised accuracy, confidentiality, and quality.^32,33^ National and international evidence shows that credentialed interpreters improve communication, reduce clinical errors, and support informed decision-making.^34,35^ Furthermore, limited interpreter access often overlaps with other structural barriers, such as digital exclusion and socioeconomic instability, thereby compounding the risk of missed care.^36^ To ensure culturally responsive care, language preferences should be documented at client intake, interpreter access should be formalised, and multilingual messaging should be integrated into recall systems. These adjustments can improve both communication quality and equity of access for individuals experiencing or at risk of homelessness.

Transport assistance was documented as a facilitator in only 3% of entries, despite consistent evidence that transport difficulties are a significant barrier to healthcare attendance among people experiencing homelessness.^37–40^ Although appointment assistance appeared in 71% of facilitator entries, 82% of recorded challenges involved appointments not being booked at the point of care. Introducing a “book-before-discharge” workflow could prevent missed recalls and ensure continuity for clients who rely on public or assisted transport.

Relocation and service disengagement disrupted continuity of care. There were no formal handover processes when clients moved or could not be reached, which risked loss to follow-up and discontinued eye and social care. Research shows that fragmented transitions between hospital and community services are a significant barrier to care for people experiencing homelessness, contributing to higher readmission rates and unmet health needs.^41–43^ In hospital systems, structured handovers and discharge planning are standard safety practices, yet equivalent continuity frameworks are rarely implemented in community or outreach settings.^44^ Introducing formal handover templates, shared care protocols, and clear points of contact between optometry, social work, and external providers could improve information continuity and follow-up, particularly for clients with unstable housing or elevated clinical risk.

Social work played a central role in bridging service gaps. Documentation highlighted assistance with transport, advocacy, appointment scheduling, and follow-up coordination. These supports addressed both practical and relational barriers to engagement. Evidence from integrated homeless health services shows that social workers are uniquely positioned to bridge organisational boundaries, coordinate follow-up, and maintain trust among clients facing complex psychosocial barriers.^45^ Their advocacy and logistical support are associated with improved continuity of care and service engagement among clients with unstable housing or limited digital access.^46,47^ These findings are consistent with broader evidence on the role of social workers in primary and community care, where they facilitate communication, integrate social and clinical information, and enhance continuity across fragmented service systems.^48^ Embedding these roles systematically within interdisciplinary teams can strengthen service cohesion, formalise shared responsibility for follow-up, and ensure that care pathways remain accessible even when clients relocate or have unstable contact details.

Overall, these service gaps point to structural rather than isolated client factors.^6,49,50^ Reliance on informal interpreters, lack of structured handover for client relocations, and frequent disconnected/unstable phone contacts reflect underinvestment in culturally responsive and continuity-focused systems. This aligns with the concept of structural competency, which interprets health inequities as outcomes of poor system design and institutional neglect rather than individual behaviour.^51^ These structural factors shape the conditions under which patient–provider interactions occur and influence who can access and sustain care. Social work documentation makes these mechanisms visible by showing how material resources (transport) and immaterial supports (information, advocacy) are provided, or withheld, and how that shapes access to care. In this context, accessibility is relational, reflecting both the health system’s capacity and clients’ ability to engage within its constraints.^52^ Operational refinements such as standardised outcome coding, redundant contact pathways, and structured appointment-booking workflows represent low-cost adjustments that could immediately improve continuity of care and recall accuracy for this population. Systematic review of these encounters should inform service improvement and guide structural changes that advance rights-based, equitable care. These operational recommendations directly enact the Triple Mandate by linking client-level barriers with organisational accountability and professional ethical standards.

Interprofessional education was an additional strength of this study. Optometry and social-work students, supervised by registered practitioners, jointly contributed to assessments and follow-up. Evidence from interprofessional education literature shows that such experiences improve teamwork, role clarity, and preparedness to work with priority populations.^53–59^ Collectively, these findings reinforce the value of embedding interprofessional education within co-designed, interdisciplinary models that integrate optometry and social work to strengthen collaboration, streamline service delivery and promote health equity for individuals experiencing or at risk of homelessness.^60–62^

Strengths of the study include including a priority framework tied to clinical and social risk, and systematic documentation of communication attempts and client-identified facilitators. This student-led design also fostered interprofessional education, preparing future clinicians to collaborate effectively in high-priority settings. Collectively, these features represent a significant step towards a more equitable and rights-based vision care. Limitations include a single-centre design over seven months, variable denominators, and reliance on routine case-note detail which may have introduced under-ascertainment or misclassification of outcomes. For example, 42% of call records were labelled as unclear or unspecified, and SMS response data were largely absent, limiting the accuracy of recall-success estimates. The absence of client-reported perspectives is another limitation. Future implementations should include mandatory outcome coding and validation checks to minimise reporting bias. The study was descriptive and did not evaluate downstream clinical outcomes; these should be explored in subsequent work.

The findings highlight several practical strategies to strengthen service continuity at minimal cost. Implementing standardised outcome coding, offering multiple contact options, ensuring interpreter access, and adopting discharge-time booking could immediately improve recall accuracy and follow-up reliability. These are structural improvements rather than behavioural interventions and align with the Triple Mandate’s emphasis on connecting client needs, organisational accountability, and professional ethics. Embedding social work within primary eye-care teams operationalises human-rights principles and advances equity goals outlined in national and international frameworks on homelessness and health inequity.

In conclusion, this interdisciplinary collaboration between optometry and social work has demonstrated the value of a rights-based model that can reduce barriers to preventive eye care and provide actionable insights for service improvement. The next step is a prospective evaluation of the model in line with integrated-care guidance. This should measure recall success, time-to-contact, interpreter uptake, equity indicators that reach across language groups, and clinical times taken to review clinically significant findings. Embedding small tests of change, such as comparing SMS-first and phone-first contact or trialling onsite translation support, can help identify strategies that are effective and cost-efficient for this population.

## Data Availability

The data that support the findings of this study are not publicly available due to privacy and ethical restrictions but are available from the corresponding author upon reasonable request, subject to approval by the relevant ethics committee.

## Acknowledgements

The authors thank all contributors and partner organisations, including community centre staff, student volunteers, Sight For All, Baptist Care SA, Zeiss, iCare, Specsavers, and the Royal Adelaide Hospital Ophthalmology Team, for their invaluable support throughout this project.

